# Should We Delay the Second COVID-19 Vaccine Dose in Order to Optimize Rollout? A Mathematical Perspective

**DOI:** 10.1101/2021.02.13.21251652

**Authors:** Soulaimane Berkane, Intissar Harizi, Abdelhamid Tayebi, Michael S. Silverman, Saverio Stranges

## Abstract

**Objectives:** With vaccination shortage persisting in many countries, adopting an optimal vaccination program is of crucial importance. Given the slow pace of vaccination campaigns globally, a very relevant and burning public health question is whether it is better to delay the second COVID-19 vaccine shot until all priority group people have received at least one shot. Currently, many countries are looking to administer a third dose (booster shot), which raises the question of how to distribute the available daily doses to maximize the effectively vaccinated population.

**Methods:** We formulate a generalized optimization problem with a total of 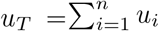 vaccine doses, that have to be optimally distributed between *n* different sub-populations, where sub-population *u*_*i*_ represents people receiving the *i*-th dose of the vaccine with efficacy *α*_*i*_. The particular case where *n* = 2 is solved first, followed by the general case of *n* dose regimen.

**Results:** In the case of a two dose regimen, if the efficacy of the second dose is less than (or equal to) twice the efficacy of the first dose, the optimal strategy to maximize the number of effectively vaccinated people is to delay the second vaccine as much as possible. Otherwise, the optimal strategy would consist of administering the second dose as quickly as possible. In the general case, the optimal vaccination strategy would be to administer the *k*−th dose corresponding to the index providing the maximum inter-dose efficacy difference (*α*_*i*_ − *α*_*i−*1_) for all possible values of *i* ∈ {1, …, *n*}, with *α*_0_ = 0.

**Conclusion:** Our results suggest that although extending the interval between doses beyond 12 weeks was likely optimal earlier in the pandemic, the reduced single dose efficacy of vaccines against the delta variant make this approach no longer viable.

## Introduction

Since December 2019, severe acute respiratory syndrome coronavirus 2 (SARS-CoV-2) has dramatically spread around the world leading to a heavy morbidity and mortality toll. The COVID-19 pandemic has put a considerable pressure on public health systems around the world with disastrous consequences on the global economy. Overcoming the COVID-19 pandemic requires massive vaccination roll out. High financial support both from private consortia and governments made it possible to develop COVID-19 vaccines extremely quickly. The production and distribution of billions of doses of COVID-19 vaccines for developing countries is the difficult challenge facing the authorities [1]. The Food and Drug Administration (FDA) has so far authorized three COVID-19 vaccines for emergency use. Two of these are mRNA vaccines (Pfizer-BioNTech and Moderna). Both of these vaccines are approved based on a regimen of two doses, although more recently a third booster dose has been recommended for high-risk persons in some developed countries after a variable duration. Initial studies by Pfizer-BioNTech demonstrated an efficacy of 95 % and Moderna has announced an initial efficacy of 94.5% after 2 doses [2]. Janssen (Johnson & Johnson) was the third COVID-19 vaccine to receive emergency use authorization. It consists on a single dose of the vaccine approved for individuals 18 years of age and older [3]. On the other hand, since December 30, 2020, ChAdOx1 nCoV-19 vaccine (AZD1222) [4], which is developed at the University of Oxford, was authorized for emergency use in the UK and then in many other countries, based on a regimen of two standard doses administered 4–12 weeks apart for adults aged 18 years and older. A single booster dose of an mRNA vaccine has also been recommended after the Janssen or ChAdOc1 vaccine at a variable interval for high risk individuals in some developed countries. Offering booster doses is not an option in most developing countries due to extreme supply shortages. Furthermore, due to the shortage in COVID-19 vaccine supplies, some countries have opted to delay the second dose of the COVID-19 vaccine for some period of time, aiming at getting the first dose of the vaccine in the arms of a large number of people, before proceeding with the second dose administration [5, 6]. This strategy has sparked some heated debates world-wide for its pros and cons, and no clear consensus is reached among experts [7, 8]. Without taking side in this matter, we tried to answer the following question, from a pure mathematical perspective: *should we delay the second dose of the vaccine or not?*. We also answer the more general question of how to optimally distribute an arbitrary number of *n* doses among the population if we adopt an *n* dose regimen. It should be noted that our model does not take into account immunological and epidemiological effects when comparing different dosing policies such as in [9, 10]. Our model can be considered primarily for general population vaccination strategies and decision making.

## Methods

First we consider the standard case of a two dose regimen. Suppose that the total number of vaccine doses to be administered daily is *u*_*T*_ = *u*_1_ + *u*_2_, where *u*_1_ is the number of first doses and *u*_2_ the number of second doses. The question that we are trying to answer is as follows: *for a fixed number of daily doses u*_*T*_, *what would be the best distribution between the number of first doses u*_1_ *and the number of second doses u*_2_ *to maximize the number of effectively vaccinated population, assuming that α*_1_(%) *is the efficacy of the first dose and α*_2_(%) *is the efficacy of the second dose*. Among the *u*_1_ population receiving the first dose, there will be (*α*_1_*/*100)*u*_1_ effectively vaccinated individuals. For those that are receiving the second dose, some of them are already effectively vaccinated due to the first dose. Therefore, among the *u*_2_ population, the number of newly effectively vaccinated individuals due to the second dose, and who were not effectively vaccinated due to the first dose, is (*α*_2_ − *α*_1_)*u*_2_*/*100. This is a consequence of the fact that the total efficacy of two doses *α*_2_ is calculated using the total number of effectively vaccinated individuals including those who are either effectively vaccinated due to the first dose or (exclusively) effectively vaccinated due to the second dose (but not the first). Therefore, the number of daily effectively vaccinated population, due to the administration of the *u*_*T*_ doses, is given by

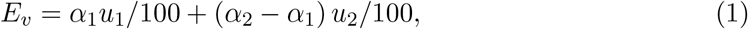

Replacing *u*_1_ by *u*_*T*_ − *u*_2_ in (1), one gets

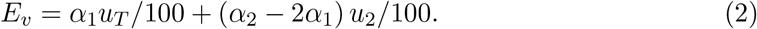

It is clear that the maximization of the daily effectively vaccinated population *E*_*v*_ depends on the sign of the term (*α*_2_ − 2*α*_1_). If (*α*_2_ − 2*α*_1_ ≤ 0), then the optimal strategy would be to set *u*_2_ = 0, and consequently the number of first doses should be equal (*u*_1_ = *u*_*T*_) which corresponds to Scenario 1. In the other case, *i*.*e*., (*α*_2_ − 2*α*_1_) *>* 0, the optimal strategy would be to set *u*_2_ = *u*_*T*_ and consequently (*u*_1_ = 0) which corresponds to Scenario 2.

### Extension to multiple doses

The above result can be generalized to a vaccination scenario with multiple doses. This is especially relevant in the current context where some regions of the world have started to administer the third dose. Experts are divided on whether it is better to start vaccination campaigns for booster doses while other people are still waiting for their first and/or second dose. Let us consider vaccination with *n* ≥ 2 dose regimen. The number of daily effectively vaccinated population, due to the administration of a total of *u*_*T*_ daily doses, is given by

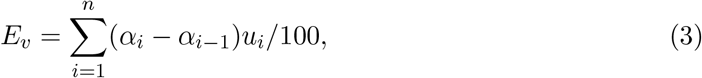

where *u*_*i*_ is the population receiving the *i*-th dose, *α*_*i*_ is the efficacy of the *i*-th dose, and *α*_0_ = 0. Note that we have the natural constraint *α*_*i*_ ≥ *α*_*i−*1_ for all *i* since the efficacy cannot decrease if another dose is administered. The goal is to maximize the linear cost function (3) subject to the following linear constraints:

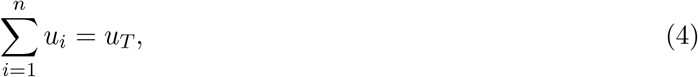

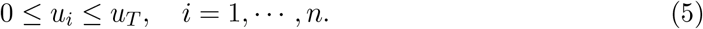

It is well-known, however, that for a linear program (LP) of this type, the maximum value of the objective function on the feasible region is one of the extreme points (vertices of the constraints polytope). Therefore, in view of the constraints (4)-(5), the optimal solution must take the following form for some *k*

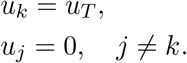

On the other hand, since the coefficients (*α*_*i*_ − *α*_*i−*1_) in the cost function (3) are all non-negative, the index *k* in the above optimal solution must be chosen such that

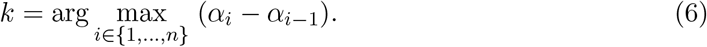

This conclusion can also be obtained using the simplex algorithm. To sum up, the optimal vaccination strategy would be to administer the *k*−th dose corresponding to the maximum inter-doses difference (*α*_*k*_ − *α*_*k−*1_) for all possible values of *k*.

## Results

In the case of a 2 dose regimen, we provide an optimal strategy for the administration of the two doses of the COVID-19 vaccine, in order to maximize the daily *effectively* vaccinated sub-population. We showed that the optimal vaccine distribution strategy is one of the following scenarios depending on the efficacy of the first and second dose. The first scenario (Scenario 1) consists of delaying the second dose of the vaccine as much as as possible. The second scenario (Scenario 2) consists of administering the second dose, as soon as possible, to those who have already taken the first dose.

We derived a simple test formula to check which of the two scenarios is optimal depending on the efficacy of the first and second doses of the vaccine. The formula is given as follows:

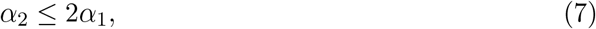

where *α*_1_ (%) and *α*_2_ (%) represent the vaccine efficacy of the first and second dose, respectively. If inequality (7) holds then we should consider Scenario 1 to obtain the best outcome of the vaccination campaign. Otherwise, Scenario 2 is preferable. An immediate consequence of this result is that if the efficacy *α*_1_ of the first dose is greater than 50%, then Scenario 1 is optimal regardless of the efficacy of the second dose since (7) must hold. This is an interesting case since most of the major vaccine companies have originally announced that their first dose of the COVID-19 vaccine is more than 50% efficient. However, there is no conclusive evidence on the real efficacy of these vaccines as studies continue throughout the world to adjust the efficacy numbers. Based on the initially announced numbers, Scenario 1 is the optimal in terms of maximizing the number of the efficiently vaccinated people per day, which is in line with the scenario adopted first by the UK authorities [6] and followed by other countries. Recent data associated with single dose vaccine efficacy in the face of the delta variant has suggested much lower efficacy with both the Pfizer and ChAdOx1 vaccines (30%) [11]. Therefore as delta has become the dominant strain globally, we now are dealing with first dose efficacy *α*_1_ that is less than 50%. In this case, Scenario 2 is optimal if the second dose efficacy exceeds the threshold 2*α*_1_, otherwise Scenario 1 is optimal. For instance if we take *α*_1_ = 40% then a second dose efficacy of more than 80% would imply that the best vaccination strategy is to provide the second dose to those who have taken the first dose as quickly as possible (Scenario 2).

On the other hand, if we consider the general scenario of *n* dose regimen, the optimal distribution of the doses between the *n* sub-populations consists in prioritizing the sub-population receiving the *k*-th dose such that (*α*_*k*_ − *α*_*k−*1_) ≥ (*α*_*i*_ − *α*_*i−*1_) for all *i* ∈ {1, …, *n*}. For example, let *n* = 3 (three dose regimen) and let *α*_1_ = 25%, *α*_2_ = 55%, and *α*_3_ = 90%. The optimal strategy, in this case, is to prioritize those taking the third dose since *α*_3_ −*α*_2_ = 35% is maximal compared to *α*_2_ − *α*_1_ = 30% and *α*_1_ − *α*_0_ = 25%.

## Discussions

Work done early in vaccine development [12] reported a vaccine efficacy of 94.8% (95% CI: 89.8–97.6) against SARS-CoV-2 after 7 days of being fully vaccinated with two doses of the messenger RNA (mRNA) vaccine BNT162b2 (Pfizer–BioNTech). In the same paper, the authors report a vaccine efficacy of 52.4% (95% CI: 29.5–68.4) from after the first dose to before the second dose. Theoretically speaking, the 52.4% first dose efficacy would imply that the optimal vaccination strategy consists in delaying the second dose. However, this conclusion is not definitive yet since the lower bound (29.5%) of the confidence interval does not satisfy the formula (7). In the recent correspondence [13], the authors pointed out that the data used in [12] to assess the first dose efficacy were collected during the first 2 weeks after the first dose, when immunity would have still been mounting. The authors have reassessed the efficacy of the first BNT162b2 dose and report 92.6% (95% CI: 69.0–98.3) from after the second week of taking the first dose to before the second dose. With this high first-dose efficacy, our theoretical findings support the fact that the benefits of the BNT162b2 vaccine could be maximized by deferring second doses until all the population (or at least the priority group members) are offered at least one dose. The same conclusion can be derived for the mRNA-1273 vaccine (Moderna) which achieved a 94.1% (95% CI: 89.3-96.8%) with a two dose regimen as reported in [14, 15] while the efficacy of the first dose after two weeks was about 92.1% (95% CI: 68.8-99.1%). On the other hand, the ChAdOx1 nCoV-19 vaccine (AZD1222), developed at the University of Oxford, is a chimpanzee adenoviral vectored vaccine with full length SARS-CoV-2 spike insert. Efficacy of two doses of the vaccine in the interim analysis [4], which pooled data from Brazil and the UK, was 70·4% (95% CI: 54.8–80·6) overall (with 4 weeks time interval between the two doses). Since December 30, 2020, ChAdOx1 was authorised for emergency use in the UK and then in many other countries, based on a regimen of two standard doses administered 4–12 weeks apart for adults aged 18 years and older. In the recent primary analysis [16], the standard regimen with two doses has shown an efficacy of 66·7% (95% CI 57·4–74·0) after 14 days of the second dose. Exploratory analyses showed that a single standard dose of ChAdOx1 nCoV-19 had an efficacy of 76·0% (95% CI 59·3-85·9) from day 22 to day 90 after vaccination [16]. Interestingly, after the second dose, efficacy was higher in those with a longer vaccination interval (vaccine efficacy 81·3% [95% CI: 60·3–91·2] at ≥ 12 weeks interval) than in those with a short interval (vaccine efficacy 55·1% [95% CI: 33·0-69·9] at *<*6 weeks) which was is predominating locally. Overall these data confirm that in the period shortly after vaccine rollout, the approaches taken by the UK and Canada to extend the interval between doses was likely optimal for population health.

There are still concerns, however, about the vaccine effectiveness in older adults and immunocompromised population [17]. The original randomized trials by which the vaccines were licensed, usually contained limited numbers of highly immunocompromised patients, but some of these trials did have large numbers of elderly patients. The efficacy of the BNT162b2 (Pfizer–BioNTech) was calculated using a trial group that contained 42.2% of people aged more than 55 years [12] while the efficacy of the Moderna vaccine was assessed based on a trial group with 24.8% of people aged more than 65 years (efficacy of the vaccine dropped by around 10% for this group) and 16.7% were younger than 65 years of age and had predisposing medical conditions that put them at risk for severe COVID-19. Astra-Zeneca was initially criticized for being an exception, with only 8% of patients over 65. On the other hand, the immunogenicity data in [16] showed binding antibody responses more than two-fold higher after an interval of 12 or more weeks compared with an interval of less than 6 weeks in those who were aged 18–55 years. This correlation, however, was not observed in those who are more than 56 years old. In fact, the efficacy of the first dose of the vaccine is most likely different in different patient populations. In the elderly and immunocompromised populations, the efficacy of one dose may be dramatically reduced compared to that of healthy young people. Since these populations are at the highest risk, it makes sense to prioritize them along with the front-line health workers by providing them with the two doses of the vaccine as soon as possible (using the standard interval). For the rest of the general population, the delivery of the second dose of the vaccine could be stretched to the maximum recommended time by health authorities if the first and second dose efficacies satisfy inequality (7).

A prioritization strategy among the general “healthy” population based on neutralizing antibody levels may also be possible once a correlation with protection against the virus is clearly established and quantified. Large population wide studies could be used to get sub-population data on (first and second dose) vaccine efficacy to inform more targeted strategies.

Most studies of delay in dose strategies assume that the initial protection afforded by a single dose will not decline at an accelerated rate when compared to a two dose vaccine schedule. Ongoing studies to confirm this will be required, as this will determine the maximum interval that vaccine can be safely delayed. In Canada a 4 month interval was initially undertaken whereas in the United Kingdom a 3 month interval was used while vaccine stocks were low and prior to the emergence of the delta variant. Ongoing real world efficacy data has been generated through these national programs. Our analysis would suggest that when the efficacy of a single dose falls below 50%, further delays in the second dose could be unwarranted. Finally, complete avoidance of the second dose may be considered for certain populations, such as those with a history of previous COVID, in whom one dose may provide full protection [18].

Although the pace of vaccination is accelerating around the world, the question of whether to delay the second dose or not is still very relevant for many countries which are struggling with severe vaccine shortages. The new issue is related to which strain of virus is predominating locally. The effectiveness of a single dose of BNT162b2 and ChAdOx1 COVID-19 vaccines is remarkably different by strain, and was reported to be only 33.5% for the Delta variant (B.1.617.2) cases and 51.1% for the Alpha variant (B.1.1.7) cases [19]. Unfortunately, recently the delta variant has spread globally to become dominant on every continent. Therefore, a single dose strategy is no longer likely to be effective. This emphasizes the importance of urgently providing enough vaccine to allow the two-dose regimen to be rolled out globally, particularly in order to prevent the emergence of more transmissible, more virulent or possibly vaccine resistant strains.

Finally, we note that the success of a vaccination campaign depends also on the level of herd immunity achieved. The herd immunity level that needs to be achieved in order to end a pandemic is tightly dependent on the basic reproduction number *R*_0_ [20]. At the early stages of the COVID-19 pandemic, the estimates of *R*_0_ ranged from 1.4 to 6.49 with an average of around *R*_0_ = 4 [21], which translates to a herd immunity level of about 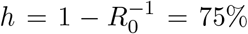. With a vaccine efficacy between 85% − 95%, we need to vaccinate at least around 79% − 88% of the population to help ending this pandemic while aiming for a quick ‘back to normal’. Unfortunately with the delta variant now predominating and having an *R*_0_ ranging between 3.2 to 8 [22], with an average of 5.6, we will need to vaccinate around 86-96% of the population.

## Data Availability

There is not data associated with this paper.

